# Attitude of reproductive age women towards male involvement in family planning; a community-based cross-sectional study in Nakawa Division, Kampala, Uganda

**DOI:** 10.1101/2022.07.14.22277630

**Authors:** Sarah Namee Wambete, Ararso Baru, Dorcas Serwaa, Edem Kojo Dzantor, Evelyn Poku-Agyemang, Margaret Wekem Kukeba, Oladapo O. Olayemi

## Abstract

**Background:** In African countries, men are often the primary decision-makers and that have a significant influence on their spouse’s health and access to health care including family planning (FP) decisions. This study aimed to assess the attitude of women in Nakawa Division, Kampala, Uganda towards male involvement in FP and the associated factors.

**Methods:** This was a community-based cross-sectional study carried out in Nakawa Division, Kampala, Uganda. A total of 480 women aged 18-49 years were selected as participants for the study using multi-stage sampling. A semi-structured interviewer-administered questionnaire was used to collect information from the participants. The data collected was analyzed using SPSS version 22. Data were described using frequency and percentage while associations were assessed using logistic regression analysis at P<0.05.

**Results:** A total of 485 participants with mean age of 28.29±6.57 years were involved in this study; 197(41.0%) were aged 26-33 years, 399 (83.1%) were Christians, 240(50.0%) had attained secondary school education and 239(49.8%) of their partners had attained tertiary education. The most utilized contraceptives among the women were injectables 151(32.5%), pills 122(26.2%), condoms 76(16.3%), implants 37(8%) and calendar method 30(6.5%). More, 302/465(62.9%) of the women had adequate partner involvement in their FP and a total of 438/480 (91.3%) of the participants had favorable attitude of women toward male involvement in FP. After adjusting for confounders, participants with an average monthly income of 600,000/= and above were more likely to have favorable attitude towards male involvement in FP (AOR=10.51, 95% CI {1.19−93.25}, p=0.035) compared with those earning 0-100,000/= average monthly income per month. Also, participants with adequate male partner involvement in FP/contraceptive use were more likely to have favorable attitude towards male involvement in FP (AOR=2.78, 95% CI {1.23−6.30, p<0.014)

**Conclusion:** The study found high favourable attitude of women towards male partner involvement at FP. The average monthly income of participants and male involvement were predictors of favourable attitude towards male involvement in FP. This finding indicates the need for increased sensitization of the men as a means of attaining the broader objective of increasing male partner involvement in FP for better contraceptive use and better birth spacing.

## Introduction

In Uganda, the total fertility rate (TFR) stands at 5.4 children per woman with those in rural areas more likely to have more children than their counterparts in urban areas [1]. Over the years, between 2011 and 2015, the Uganda national contraceptive prevalence rate (CPR) has gone from 24% to 39% and the use of modern contraception is higher among women in urban areas (41%) than those in rural areas (33%) [1]. The progress is still very poor and slow and the figures are still quite alarming [1, 2]. An array of circumstances may influence women’s decision to use FP methods including knowledge, attitude, accessibility and affordability to health facilities/family planning clinics [3, 4]. Regardless, the role of men in FP uptake cannot be underestimated as studies have shown that, marriage and cohabitation harm modern contraceptive use [5–7].

Is a common knowledge that FP is regarded as a woman’s “thing” in most African countries. However, in the African setting, men play an important role in family decision-making and the health seeking behaviour of the family including the uptake of available FP services [8]. Studies continue to demonstrate the importance of men in reproductive health decisions and improvement in the utilization of FP services in both women and men [9, 10]. Previous studies have shown that, FP programs that target couples have proved to be more effective than those directed to individuals either male or female [9, 11]. With men’s significant influence in family and health related seeking behaviours and FP service uptake especially among women; the important question to ask is how are women feeling about male’s involvement their (women) use of FP? Some studies have reported that in some cases, males involvement in FP decisions but deter their partners from using FP methods and such negative attitude and heavy resistance of men might contribute to low utilization of FP methods in women [12, 13].

To strengthen male involvement in sexual and reproductive health (SRH) in Uganda, Reproductive Health Uganda (RHU) and the Ministry of Health Uganda (MHU) in partnerships with local stakeholders, have been carrying out activities that involves men as agents of change, equal partners and clients. For example, gender-focused discussions and male-only information and testing days are available to men at local clinics [2, 14]. Despite all these efforts, the prevalence of male involvement in SRH issues has remained significantly low.[15–17] Most descriptive studies aimed at ascertaining male involvement in FP has involved men whiles women’s attitudes towards male involvement has been neglected. Therefore, this study assessed the attitude of women in Nakawa Division, Kampala, Uganda regarding male involvement in FP and the associated factors.

## Subjects and Methods

### Study Area and population

The study was conducted in Nakawa division, the largest among the five administrative divisions of the city of Kampala district in Uganda bordering Kira Town to the east, Wakiso District to the north, Kawempe Division to the north-west, Kampala Central Division to the west and Makindye Division across the south. The division which is located in the eastern part of Kampala city with coordinates 0°20’00.0”N, 32°37’00.0”E. The 2019 national census estimated the division’s population at 317,023, with 163,594 females and 153,429 males. The population growth rate in 2019 was 4.8 percent, the total fertility rate was 5.1 percent and the maternal mortality rate was 116 per 100,000 live births [18].

### Study design, sample size estimation and sampling

This community-based cross-sectional study carried out in Nakawa Division, Kampala, Uganda from November, 2018-June 2019. The sample size for the study was calculated using the single population proportion formula and based on both partner involvement in contraceptive decision rate of 24%, from previous study conducted in Uganda [19], with a 5% margin of error and a confidence interval of 95%; given that Z (at 95% confidence interval) =1.96 and considering 15% for non-response rate, the calculated sample size including was 320. However, because multi-stage sampling was used, a multiplier factor of 1.5 was considered to the sample to cater for the design effect to give a final sample of 480. Therefore, a total of 480 women aged 18-49 years were recruited in this study.

Multistage sampling technique was employed to select study participants. Of the 23 parishes in Nakawa division, eight perishes namely, Banda, Bukoto I, Butabika, Kiswa, Kiwatule, Mutungo, Naguru I and Naguru II were selected by random sampling. The sampling frame was prepared for each perish after identifying households with women aged 18-49 years through rapid registration and list development during house-house visit by the trained village health teams. The sample size was distributed to each of the selected parish proportional to their population size (women aged 18-49 years). The sampling interval was determined by dividing the total population in each parish to the size of the sample required from that parish and participants were selected using systematic random sampling. The phone numbers of consenting eligible participants were requested for further discussions.

### Study instrument and data collection

A close-ended structured questionnaire was used to collect information from the participants. The questionnaire consisted of three sections namely section A: sociodemographic and economic characteristics of the participants and section B: questions designed to measure male involvement in family planning such as (a) the discussion of family planning with partner, (b) financial support for family planning from partner, (c) joint decisions with partner about contraceptive use and choice, (d) attendance to family planning clinics at least once with the past 12 months. Section C consisted of an index of six (06) questions aimed at measuring women’s attitudes towards male involvement in FP & CU as favourable or unfavourable.

The eligible participants were contacted two weeks prior to the data collection and the purpose, risk and significance of the study was introduced to them. Participants who met the inclusion criteria were mobilized to the nearest community center for data collection for data collection. Before the questionnaires were administered, the aim and objectives were explained to participants and their consent was then sought. Participants were informed of their right to decline to partake in the study and confidentiality was assured and kept. Rapport was established and data was collected using the structured interview-administered questionnaire. The face-to face survey were conducted by trained research assistants with minimum qualification of a university diploma, who were fluent in both Luganda and English. From time to time, the participants were interacted with by repeating or clarifying questionnaire items to encourage prompt responses from the participants. The survey was estimated to take approximately 10-15 minutes to complete.

### Selection Operational Definitions

To determine the overall male involvement in FP, participants were asked whether their spouse/partner performed a specific single task and scored (1) for where a task was performed and (0) for where the task was not performed. The overall score a participant could obtain ranged from 0 to 5. Therefore, a score of 3 and above indicated adequate male involvement and a score of 2 tasks and below indicated inadequate male involvement. With regards to attitude of women towards male involvement in FP, six (06) questions with dichotomous (Agree and Disagree) with answers were asked. A maximum score a participant could obtain was 6 and a minimum of 0, a participant was presumed to have a favourable attitude on male involvement in FP & CU if she scored three or more points otherwise had an unfavourable attitude.

### Ethical Consideration and informed consent

This study was conducted based on the Helsinki Declaration and study protocol, consent forms and participant information material were reviewed and approved by University of Ibadan/University Collage Hospital Ethics Committee (UI/EC/18/0635). Ugandan National Council for Science and Technology approved the research and assigned a reference number (UNCST: SS 4921)

Written informed consent of individual participants was sought after the aims and objectives of the study had been thoroughly explained to them. Participants either signed or thumb-printed to give their consent, before the commencement of the study and they assured of the confidentiality of their data.

### Statistical Analysis

After retrieving the questionnaires, each was inspected for accuracy and completeness. Data entry, and analysis was done using SPSS software (version 22). The sociodemographic and economic characteristics, male involvement and attitude of the participants were analysed and presented as frequencies, percentage, mean, standard deviations and presented appropriately using tables. Binary and multivariate logistic regression analysis was applied to explore the association between the women’s attitude towards male involvement in FP and participants characteristics. A p value <0.05 was considered statistically significant.

## Results

### Socio-demographic and economic characteristics of the participants and their partners

Table 1 depicts the main sociodemographic characteristics of the study participants. A total of 485 participants with mean age of 28.29±6.57 years were involved in this study; 197(41.0%) were aged 26-33 years, 399 (83.1%) were Christians, 240(50.0%) had attained secondary school education and 239(49.8%) of their partners had attained tertiary education. The majority of the participants and their partners were married (361,75.2% vs 431,89.8%), lived in urban area (342,71.3%), and employed (381,79.4%). More than two-thirds of participants and their partners worked as full time (342, (71.3%) vs 382(79.6%)). Also, 205(42.7%) of the participants earned between 120,000-500,000 UGX on average per month and most 382(79.6%) of their partners earned 600,000 UGX and above.

**Table 1:** Socio-demographic and economic characteristics of the participants and their partners.

| Variables | Frequency (N=480) | Percentage (%) |
| --- | --- | --- |
| **Age of participants (yrs)** | Min=18 Max=49 | Mean: 28.29±6.57 |
| 18-25 | 192 | 40.0 |
| 26-33 | 197 | 41.0 |
| 34-41 | 63 | 13.1 |
| 42-49 | 28 | 5.8 |
| **Religion** |  |  |
| Christian | 399 | 83.1 |
| Muslim | 81 | 16.9 |
| **Education level of participants** |  |  |
| No formal education | 18 | 3.8 |
| Primary | 62 | 12.9 |
| Secondary | 240 | 50.0 |
| Tertiary | 160 | 33.3 |
| **Education level of partners** |  |  |
| No formal education | 43 | 9.0 |
| Primary | 24 | 5.0 |
| Secondary | 174 | 36.3 |
| Tertiary | 239 | 49.8 |
| **Marital status** |  |  |
| Never married | 96 | 20.0 |
| Ever married | 23 | 4.8 |
| Married | 361 | 75.2 |
| **Area of residence** |  |  |
| Rural | 21 | 4.4 |
| Semi-urban | 117 | 24.4 |
| Urban | 342 | 71.3 |
| **Employment status of the participants** |  |  |
| Unemployed | 99 | 20.6 |
| Employed | 381 | 79.4 |
| **Employment status of partner** |  |  |
| Unemployed | 49 | 10.2 |
| Employed | 431 | 89.8 |
| **Work schedule of participants** |  |  |
| Unemployed | 99 | 20.6 |
| Shifts schedule | 7 | 1.5 |
| Part-time | 32 | 6.7 |
| Full-time | 342 | 71.3 |
| **Work schedule of partners** |  |  |
| Unemployed | 49 | 10.2 |
| Shifts schedule | 14 | 2.9 |
| Part-time | 35 | 7.3 |
| Full-time | 382 | 79.6 |
| **Average monthly income of the participants** |  |  |
| 0-100,000/= | 161 | 23.5 |
| 120,000-500,000/= | 205 | 42.7 |
| 600,000 and above | 114 | 23.8 |
| **Average monthly income of partners** |  |  |
| 0-100,000/= | 72 | 15.0 |
| 120,000-500,000/= | 176 | 36.7 |
| 600,000 and above | 232 | 48.3 |

**Table 1:** Distribution of women’s attitude toward male involvement in family planning.

| Variable | Characteristic | Frequency<br>(N=480) | Percentage<br>(%) |
| --- | --- | --- | --- |
| It is okay to discuss family planning and contraceptive use with your partner | Agree | 387 | 80.6 |
|  | Disagree | 93 | 19.4 |
| It is okay for you to attend Family planning clinics with your partner | Agree | 380 | 79.2 |
|  | Disagree | 100 | 20.8 |
| Family and friends do not mind if you discuss FP and CU with your spouse | Agree | 410 | 85.4 |
|  | Disagree | 70 | 14.6 |
| Men have a role to play in family planning and contraceptive use | Agree | 449 | 93.5 |
|  | Disagree | 31 | 6.5 |
| Discussing FP & CU with your partner motivates utilization of the services | Agree | 427 | 89.0 |
|  | Disagree | 53 | 11.0 |
| Your partner will be comfortable attending family planning sessions with you | Agree | 239 | 49.8 |
|  | Disagree | 241 | 50.2 |
| **Overall attitude** |  |  |  |
| Unfavourable |  | 42 | 8.8 |
| Favourable |  | 438 | 91.3 |

### Uptake of family planning services among the study participants

Participants were asked whether they had used any contraceptive within previous 12 months before the commencement of this study. Among 480 participants, 465 (96.8%) had used a family planning method twelve months before the study and 15 (3.2%) participants were found not to be sexually active and not using any contraceptive or not to have used one the previous 12 months before the study. Figure 1 show that, the top five most utilized contraceptives were injectables 151(32.5%), followed by Pills 122(26.2%), condoms 76(16.3%), Implants 37(8%) and calendar method 30(6.5%) (Fig. 1).

**Fig. 1:**
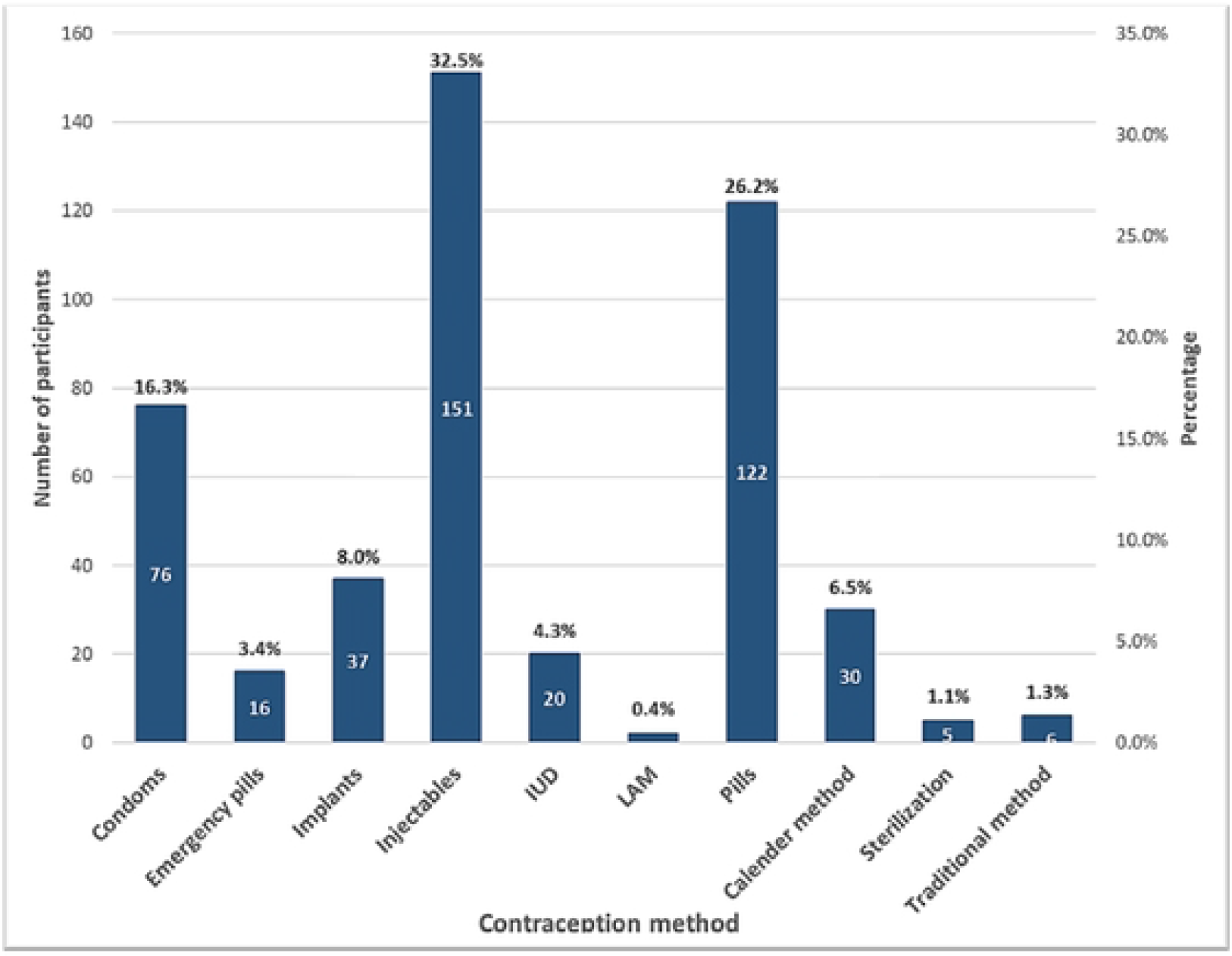
Levels of uptake of family planning services among women aged 18-49 years in Nakawa division, Kampala district, Uganda. (n=465)

### Health facility-related characteristics of participants

Among the 465/480 (96.9%) participants who were using a family planning method at the time of the study, more than half 266(57.2%) used public means of transport to the health facility to obtain their choice FP service, just over a third 173(37.2%) walked to the facility. Also, more than two-third 333(71.6%) reported living within less than five kilometres of a health facility. The majority reported a waiting time of less than an hour at the facility alone (384, 82.6%) or with partner (452, 97.2%). Almost all participants 429(92.3%) reported that the FP clinic environment is male friendly and that the behaviour of staff at the health facility was friendly 383(82.4%) (Table 2).

**Table 2:** Distribution of health facility-related characteristics of women aged 18-49 years in Nakawa division, Kampala district, Uganda.

| Variables | Frequency (N=450) | Percentage (%) |
| --- | --- | --- |
| **Transport to the health facility to obtain FP services** |  |  |
| By foot | 173 | 37.2 |
| Personal means | 26 | 5.6 |
| Public means | 266 | 57.2 |
| **Proximity to a health facility** |  |  |
| >5km | 333 | 71.6 |
| ≥5km | 132 | 28.4 |
| **Waiting time at the health facility alone** |  |  |
| >1hr | 384 | 82.6 |
| ≥1hr | 81 | 17.4 |
| **Waiting time at the health facility with partner** |  |  |
| >1hr | 452 | 97.2 |
| ≥1hr | 13 | 2.8 |
| **Rating for waiting time** |  |  |
| Short | 38 | 8.2 |
| Normal | 339 | 72.9 |
| Long | 88 | 18.9 |
| **The FP clinic environment is welcoming for men** |  |  |
| No | 36 | 7.7 |
| Yes | 429 | 92.3 |
| **The behaviour of staff at the health facility** |  |  |
| Unfriendly | 82 | 17.6 |
| Friendly | 383 | 82.4 |

### Male partner involvement in family planning use among the study participants

Table 3 summarizes the participants recall of their partners involvement in their previous FP or CU. A total of 274/465(58.9%) of the women discussed FP and CU with their partners, 342/465 (53.5%) received financial support from their partners and 251/465 (54.0%) makes joint decisions with the partner about contraceptive use and choice. In the past 12 months, less than one-third (106/465,22.8%) of the women had their partners accompany them FP clinic at least once and 329/465 (70.8%) of the participants stated that their partners were aware of their contraceptive use within the past 12 months. In totality, 302/465(62.9%) of the women had adequate partner involvement in their family planning and contraceptive use while 163/465(34.0%) had inadequate partner involvement.

**Table 3:** Distribution of male partner involvement in family planning and Contraceptive Use in women aged 18-49 years in Nakawa Division, Kampala District.

| Variables | Frequency(N=465) | Percentage (%) |
| --- | --- | --- |
| **Discusses with the partner about FP & CU** |  |  |
| No | 191 | 41.1 |
| Yes | 274 | 58.9 |
| **Partner offers financial support** |  |  |
| No | 123 | 26.5 |
| Yes | 342 | 73.5 |
| **Makes joint decisions with the partner about contraceptive use and choice** |  |  |
| No | 214 | 46.0 |
| Yes | 251 | 54.0 |
| **Partner attended FP clinic at least once in the past 12 months (n=465)** |  |  |
| No | 359 | 77.2 |
| Yes | 106 | 22.8 |
| **Partner is aware of my previous contraceptive use (in the past 12 months)** |  |  |
| No | 136 | 29.2 |
| Yes | 329 | 70.8 |
| **Overall male involvement** |  |  |
| Inadequate | 163 | 34.0 |
| Adequate | 302 | 62.9 |

### Women’s attitude towards male involvement in family planning

Table 4 summary depicts that more than three-quarters of participants agreed that it is okay; to discuss FP & CU with a partner 387/480 (80.6%), to attend FP clinics with a partner 380/480 (79.2%), that family and friends do not mind if they discussed FP & CU with a spouse 410/480 (85.4%), men have a role to play in FP & CU use 449/480 (93.5%) and to discuss FP & CU with a partner motivates utilization of the services 427/480 (89.0%). However, less than half 239/480 (49.8%) agreed that their partner would be comfortable attending family planning sessions with them. A total of 438/480 (91.3%) of the participants had favourable attitude of women toward male involvement in FP while only 42/480 (8.8%) had unfavourable attitude of women toward male involvement in FP.

### Analysis of predictors of favorable attitude of women toward male involvement in FP

Exactly 9 independent variables were entered into the bivariate and the multivariate logistic regression, the unadjusted regression analysis showed that level of education of participants, average monthly income of participants, average monthly income of partners, waiting time rating, male friendly FP clinic environment, the behaviour of staff at the health facility and male involvement were predictors of favourable attitude towards male involvement in FP. After adjusting for confounders in the multivariate analysis, only average monthly income of participants and male involvement were predictors of favourable attitude towards male involvement in FP.

The unadjusted regression analysis showed that, women who had attained tertiary education had greater odds of showing favourable attitude towards male involvement in FP compared to their counterparts who had no formal education (COR=8.85, 95% CI {2.13−36.79}, p=0.003). Participants with an average monthly income of 120,000-500,000/= (COR=1.94, 95% CI {1.00−3.75}, p=0.049) and having an average monthly income of 600,000/= and above (COR=19.8, 95% CI {2.6−148.60}, p=0.004) were more likely to have favourable attitude towards male involvement in FP compared with those earning 0-100,000/= average monthly income per month. Also, participants whose partners earned an average monthly income of 600,000/= per month had greater likelihood of having favourable attitude towards male involvement in FP (COR=2.87, 95% CI {1.14−7.23}, p=0.025). A statistically significant predictive relationship was found between favourable attitude towards male involvement in FP and women who agreed that the FP clinic environment is welcoming for men (COR=3.94, 95% CI {1.65−9.42}, p=0.002). Participants who agreed that the behaviour of staff at the health facility was friendly were less likely to have favourable attitude towards male involvement in FP (COR= 0.12, 95% CI {0.02−0.88}, p=0.037). Women who had adequate male partner involvement had higher likelihood of having favourable attitude of toward male involvement in FP (COR=3.84, 95% CI {1.90−7.77, p<0.001) relative to those with inadequate male partner involvement.

After adjusting for confounders, participants with an average monthly income of 600,000/= and above were more likely to have favourable attitude towards male involvement in FP (AOR=10.51, 95% CI {1.19−93.25}, p=0.035) compared with those earning 0-100,000/= average monthly income per month. Also, participants with adequate male partner involvement in FP/CU use were more likely to have favourable attitude towards male involvement in FP (AOR=2.78, 95% CI {1.23−6.30, p<0.014) (Table 5).

**Table 5:** Bivariate analysis of predictors of adequate male involvement in family planning among women aged 18-49 years in Nakawa Division, Kampala District, Uganda.

| Variables | Attitude |  | COR 95% CI | P-value | AOR 95% CI | P-value |
| --- | --- | --- | --- | --- | --- | --- |
|  | Unfavourable | Favourable |  |  |  |  |
|  | N (%) | N (%) |  |  |  |  |
| Women’s level of education |  |  |  |  |  |  |
| No formal education | 4(9.5) | 14(3.3) | 1 |  | 1 |  |
| Primary | 11(26.2) | 51(11.6) | 1.33[0.37–4.80] | 0.669 | 0.69[0.16–2.93] | 0.615 |
| Secondary | 22(52.4) | 218(49.8) | 2.83[0.86–9.35] | 0.088 | 1.33[0.34–5.17] | 0.683 |
| Tertiary | 5(11.9) | 155(35.4) | 8.85[2.13–36.79] | **0.003** | 4.52[0.60–34.05] | 0.144 |
| Partners’ level of education |  |  |  |  |  |  |
| No formal education | 5(11.9) | 38(8.7) | 1 |  | 1 |  |
| Primary | 7(16.7) | 17(5.0) | 0.32[0.09–1.15] | 0.081 | 0.36[0.07–1.86] | 1.859 |
| Secondary | 18(42.9) | 156(35.6) | 1.14[0.40–3.27] | 0.807 | 1.16[0.29–4.67] | 4.666 |
| Tertiary | 12(28.6) | 227(51.8) | 2.49[0.83–7.47] | 0.104 | 1.18[0.26–5.28] | 5.277 |
| Area of residence |  |  |  |  |  |  |
| Rural | 1(2.4) | 20(4.6) | 1 |  | 1 |  |
| Semi-urban | 20(47.6) | 97(22.1) | 0.24[0.03–1.91] | 0.179 | 0.25[0.03–2.47] | 0.235 |
| Urban | 21(50.0) | 321(73.3) | 0.80[0.10–5.97] | 0.798 | 0.44[0.05–4.19] | 0.475 |
| Average monthly income of the participants |  |  |  |  |  |  |
| 0-100,000/= | 24(57.1) | 135(31.3) | 1 |  | 1 |  |
| 120,000-500,000/= | 17(40.5) | 188(42.9) | 1.94[1.00–3.75] | **0.049** | 1.60[0.69–3.72] | 0.277 |
| 600,000 and above | 1(2.4) | 113(25.8) | 19.8[2.6–148.60] | **0.004** | 10.51[1.19–93.25] | **0.035\*** |
| Average monthly income of partners |  |  |  |  |  |  |
| 0-100,000/= | 9(21.4) | 63(14.4) | 1 |  | 1 |  |
| 120,000-500,000/= | 22(52.4) | 154(35.2) | 1[0.44–2.91] | 1.000 | 1.00[0.32–3.06] | 0.983 |
| 600,000 and above | 11(26.2) | 221(50.5) | 2.87[1.14–7.23] | **0.025** | 1.20[0.32–4.44] | 0.790 |
| Rating for waiting time |  |  |  |  |  |  |
| Short | 6(16.2) | 32(7.5) | 1 |  | 1 |  |
| Normal | 25(67.6) | 314(73.4) | 2.36[0.9–6.17] | 0.081 | 2.27[0.72–7.20] | 0.164 |
| Long | 6(16.2) | 82(19.2) | 2.56[0.77–8.53] | 0.125 | 1.93[0.49–7.70] | 0.349 |
| The FP clinic environment is welcoming for men |  |  |  |  |  |  |
| No | 8(21.6) | 28(6.5) | 1 |  | 1 |  |
| Yes | 29(78.4) | 400(93.5) | 3.94[1.65–9.42] | **0.002** | 2.40[0.85–6.78] | 0.098 |
| **The behaviour of staff at the health facility** |  |  |  |  |  |  |
| Unfriendly | 1(2.7) | 81(18.9) | 1 |  | 1 |  |
| Friendly | 36(97.3) | 347(81.1) | 0.12[0.02–0.88] | **0.037** | 0.16[0.02–1.22] | 0.076 |
| **Male involvement** |  |  |  |  |  |  |
| Inadequate | 24(64.9) | 139(32.5) | 1 |  | 1 |  |
| Adequate | 13(35.1) | 289(67.5) | 3.84[1.90–7.77] | **<0.001** | 2.78[1.23–6.30] | **0.014\*** |

## Discussion

Our study assessed the attitudes of women towards male involvement in FP and the associated factors. The mean age of the participants was 28.29±6.57 years, signifying a youthful population and a reflection of Ugandan’s rural and urban women’s age structure where most are in their second and third decades of life [20]. This stresses a need to control fertility rates through the promotion of FP services among the population. Consistent with the findings of Andi et al. [5] most of the participants were Christians and with secondary education. These findings are important for public health interventions planning. Leaders of the Christian religion could be considered as key stakeholders in the planning and promotion of FP services in Nakawa Division, and across the country. They can use their platforms to promote the use of FP among the population. It is equally important to recognize other minority religious groups in addressing FP in Uganda since studies elsewhere have described the role of religion in the promotion of contraceptive use [21, 22].

According to the Ugandan Demographic and Health Survey (UDHS) [20] persons with secondary education are considered literates. Therefore, having majority of the current study’s participants and other studies [5, 23, 24] with secondary and postsecondary education is important for increased health literacy on FP. Considering the significant number persons who enrolled to the secondary level of education, it is important to strengthen the effectiveness of the implementation of the Ugandan School Health Policy[25], with much emphasis on FP services and importance. Majority of the participants were married, highlighting the need to consider partners in FP interventions [26]. The study observed that most of the participants were urban dwellers and was inconsistent with the finding of Andi et al. [5]. Suggestively, interventions on FP should be targeted among both urban and rural areas of the country. For example, the UDHS [20] reported that women in the urban areas are more literate than women in rural areas of Uganda. Therefore, public health education on FP should be targeted across the country with more concentration at the rural areas.

Contraceptive uptake was significant among participants in our study. Contraceptive uptake in Uganda is universal [23, 24, 27]. For example, Otim [24] reported varying but significant proportions of contraceptive use among Ugandan women with the Central, Eastern, Northern and Western parts of the country recording 65%, 57%, 45% and 56% contraceptive use respectively. Though there is universal uptake of contraceptives in our study setting and across the country, persons who do not use available contraceptives equally form important section of Uganda’s population. Such persons should be targeted for FP services and interventions to address the unmet needs of FP in the country. Consistent with previous studies in Uganda [19, 28, 29] the commonly utilized contraceptives comprised injectables, pills, condoms, implants and calendar methods.

To access available FP services, most of the participants used public means of transport with majority dwelling in catchment areas of FP. Turnaround time for FP services was less than an hour for most users of FP and more importantly male friendly. Though most of the participants reside in the catchment area for FP services, there is a need to make FP more accessible to the 28.4% who are distant from FP service centres. A previous study in Uganda reported some barriers to FP services use to comprised long distance to health facility and husbands been unfriendly towards the use of FP [30, 31]. The apparently short distance and male friendly FP service environment in our study suggests an improvement in accessibility to FP services in the study. Most of our participants self-reported adequate involvement of their male partners in their patronage of FP services and consistent with studies elsewhere [32, 33]. In the African traditional setting, males play important role in the decision-making and the health seeking behaviours of the family [33].

Our study participants indicated that their partners’ involvement in their FP usage comprised having discussions on FP choices, financial support, and accompaniment to FP service centres. Therefore, to scale-up FP services uptake in Uganda, males should be targeted for public health educational and promotional activities to increase their appreciation of FP services. The male’s partner appreciation of FP services could go a long way to encourage female patronage of FP services. Mulatu et al. [34] reported that male’s involvement in discussions on FP, sexual and reproductive health, and approval from husbands on the use of FP were positive factors for FP use.

On participants’ attitudes towards male involvement in FP services, our study showed that most of the women had favourable attitudes towards males’ involvement in FP services. Another important assertion made by participants was feeling comfortable to discuss FP services with their male partners. Previous studies have extensively explored the importance of spousal communication and its’ effect on the use of FP services including contraceptives [33, 35]. Zelalem et al. [35] reported that effective spousal FP communication was significantly associated with current modern contraceptive use.

Key predictors for favourable attitude towards male involvement in FP after adjusting for confounders in the multivariate analysis included average monthly income of participants and male involvement of FP. Our finding of average income as a positive predictor for FP attitude was consistent with Serwanja et al. [28] report of being in the middle wealth quintile as a positive factor for contraception. Similarly, Our finding of male involvement as a positive factor for favourable attitude towards male involvement in FP was inconsistent with Asif et al. [36] finding of women with partners who discourage usage of contraception. Though there exist variations in the predictors for FP services for male involvement, which is a common phenomenon in research due to factors including variations in sample sizes and study context.

Our results indicate that income and male involvement in FP services were important factors to determine female attitudes towards FP services. Income has been reported by previous studies and reports as a the most important social determinant of health. An individual’s level of income influences the general living conditions, including the health-related behaviours [37, 38]. Our study may suggest that males that show financial support to their spouses are likely to afford FP planning services. Again, the finding highlight the need for the Ugandan government to address the poverty gap for improved health seeking behaviours including uptake of FP services.

This study was without limitations. Frist. the study was self-reported and may be subjected to information bias. Second, the study reports on males’ involvement on family planning from the females and may not be reflection of male involvement in family planning services. The study is also limited to the Nakawa Division, Kampala, and may not reflect the population of Uganda. Therefore, the use of the study for decision making purposes on family planning should be done with caution and contextualized.

## Conclusion

Family planning uptake was high among participants and their male partners. Common utilized contraceptives were injectables, pills, condoms, Implants and calendar method. The overall attitudes of women towards partner involvement in their family planning and contraceptive use was adequate. Average monthly income and participants with adequate male partner involvement in family planning use were more likely to have favorable attitude towards male involvement in FP. The study shows the need to involve males in family planning programmes for increased service uptake.

## Data Availability

All data and related metadata underlying the findings reported are provided as part of the submitted article

## Acknowledgements

Our deepest gratitude goes to Pan African University, University of Ibadan, Ugandan National Council for Science and Technology, The District Health Office of Nakawa Division and Kampala Capital City Authority for facilitating this study. Finally, our special respect goes to all respondents and data collectors in this study.

## Authors’ Contributions

SNW made substantial contributions to conception, design, acquisition of data and assisted by AB and OOO. Data analysis and interpretation was done by AR, DS and EKD. SNW, EP, MWK and OOO wrote the first draft of the manuscript. All authors critically reviewed, revised and approved the final manuscript

